# SARS-CoV-2 Reinfection in Patients Negative for Immunoglobulin G Following Recovery from COVID-19

**DOI:** 10.1101/2020.11.20.20234385

**Authors:** Ayad M. Ali, Kameran Mohammed Ali, Mohammed Hassan Fatah, Hassan Mohammad Tawfeeq, Hassan Muhammad Rostam

## Abstract

While many patients infected by severe acute respiratory syndrome coronavirus 2 (SARS-CoV-2) eventually produce neutralising antibodies, the degree of susceptibility of previously infected individuals to reinfection by SARS-CoV-2 is currently unknown. To better understand the impact of the immunoglobulin (IgG) level on reinfection in recovered coronavirus disease 2019 (COVID-19) patients, IgG levels against SARS-CoV-2 were measured in 829 patients with previously confirmed infection just after their recovery. Notably, 87 of these patients had no detectable IgG concentration. While there was just one case of asymptomatic reinfection 4.5 months after the initial recovery amongst patients with detectable IgG levels, 25 of the 87 patients negative for IgG were reinfected within one to three months after their first infection. Therefore, patients who recover from COVID-19 with no detectable IgG concentration appear to remain more susceptible to reinfection by SARS-CoV-2, with no apparent immunity. Also, although our results suggest the chance is lower, the possibility for recovered patients with positive IgG findings to be reinfected similarly exists.

## Introduction

Coronavirus disease 2019 (COVID-19) is an infectious disease caused by a novel coronavirus, severe acute respiratory syndrome coronavirus 2 (SARS-CoV-2), which was named so given the similarity of its symptoms to those induced by severe acute respiratory syndrome [1]. Since the first reports of a viral pneumonia of unknown origin emerged from China in late 2019, this disease has spread across the world, with new cases reported daily. The clinical manifestations of COVID-19 range widely from asymptomatic to mild, moderate and rapidly progressive severe (pneumonia) disease that can lead to death in some individuals [2-4]. The moderate clinical symptoms of patients with COVID-19 include fever, dyspnoea, fatigue, dry cough, myalgia and pneumonia. In severe cases, affected patients may experience acute respiratory failure, septic shock and organ failure that might culminate in death [5, 6].

Transmission of SARS-CoV-2 from infected people to others is suggested based on epidemiology and clinical evidence [7, 8], with even asymptomatic infected individuals believed to be capable of transmitting the virus [9, 10].

Infection by SARS-CoV-2 leads to a detectable immune response, but the susceptibility of previously infected individuals to reinfection by SARS-CoV-2 is not well understood given the brevity of the worldwide pandemic to date. Generally, infection results in the generation of neutralising antibodies in patients [11] [12]. SARS-CoV2 has the capacity to escape innate immune responses, which allows the pathogen to produce large numbers of copies in primarily infected tissues, usually airway epithelia [13]. Principally, patients who recover from infectious diseases are usually immunised henceforth against infection by the causative virus; however, reinfection by respiratory viruses is extremely common among humans of all ages due to these viruses’ progressive evolution through RNA genome mutations that lead to antigenic drift and immune escape. However, the complete mechanisms governing our susceptibility to recurrent viral infections remain poorly understood [14, 15]. Although some studies indicate the persistence of protective immunoglobulin IgG levels in the blood, saliva and other body fluids for months after infection with SARS-CoV-2 [16, 17], limited numbers of case studies of patients with COVID-19 have reported positive test results after the disease symptoms had resolved and negative test results were recorded, supporting the possibility of reinfection [18-20]. These reports included both patients with mild disease [21, 22]and others with more severe conditions [20, 23].

This study aimed to report an additional group of COVID-19 patients who were reinfected by SARS-CoV-2 and argue that the IgG level is a potential marker of the reinfection risk.

## Materials and Methods

### Study population

The study included a group of 829 patients admitted to Qala Hospital, Kalar, Kurdistan region, Iraq from the last week of May until the middle of October.

### Real-time reverse-transcription polymerase chain reaction (RT-PCR) assay for the diagnosis of SARS-CoV-2

Pharyngeal swabs were administered to extract SARS-CoV-2 RNA from each patient; then, the total RNA was extracted using the AddPrep Viral Nucleic Acid Extraction Kit (Addbio Inc., Daejeon, South Korea). Next, The presence of the SARS-CoV-2 virus was detected by real-time RT-PCR amplification of the SARS-CoV-2 open reading frame 1ab (ORF1ab) and envelope (E) gene fragments. The amplification reactions were carried out with 10 µL of 2X RT-PCR mastermix, 5 µL of primer/probe mix and 5 µL of template RNA for a final volume of 20 µL using the PowerChek SARS-CoV-2 Real-time PCR Kit (Kogenebiotech, Seoul, Korea), described previously [24]. We followed the kit’s instructions and adopted the following thermocycler protocol: 50°C for 30 minutes and 95°C for 10 minutes, followed by 40 cycles of 95°C for 15 seconds and 60°C for one minute. When findings regarding the two target genes (ORF1ab, E) were positive according to specific real-time RT-PCR, a sample was defined as positive if the viral genome was detected at the cycle threshold value (Ct-value) of 36.7 or less, while the Ct-value of greater 36.7 was defined as indicating a negative test result or recovery (i.e., disappearance of signs and symptoms in a previously RT-PCR positive patient).

### Enzyme-linked Immunosorbent Assay

A serum sample was collected from patients with a confirmed SARS-CoV-2 RT-PCR test result just after their recovery.

The anti–SARS-CoV-2 IgG antibody level was assessed using a commercially available SARS-CoV-2 IgG test kit (Pishtaz Teb Diagnostics, Tehran, Iran) targeting the nucleocapsid (N) antigen of the SARS-CoV-2 virus. Sera were diluted 1:101. First, 10 µL of the specimen together with 1,000 µL of the sample diluent was processed in a 96-well test kit. Then, 100 µL of each control serum and diluted specimens were placed into the appropriate well, with the first two wells chosen as blanks and the next two chosen as negative control wells. Positive controls were used as duplicates and the other wells were used for samples. Based on the manufacturer’s formula; the following cutoffs were applied: 1.1, positive; 0.9 to 1.1, equivocal; and less than 0.9, negative.

### Ethics declarations

All methods were used in accordance with relevant guidelines and regulations. Also, we confirmed that all experimental protocols were approved by the Ethics Licensing Committee of the Kalar Technical Institute at the Sulaimani Polytechnic University (no. 02 on 01/08/2020). In addition, informed consent was obtained from all study participants or a parent and/or legal guardian if the individual was younger than 18 years of age.

## Results

Our study found that 87 patients tested negative for IgG specific to SARS-CoV-2 after recovery among a population of 829 patients who were infected with SARS-CoV-2 for the first time. Twenty-six patients (14 male and 12 female patients, aged 10–60 years old) were reinfected after recovery; of these, 25 patients were in the IgG-negative group and only one patient was IgG-positive (Figure 1).

**Figure 1:**
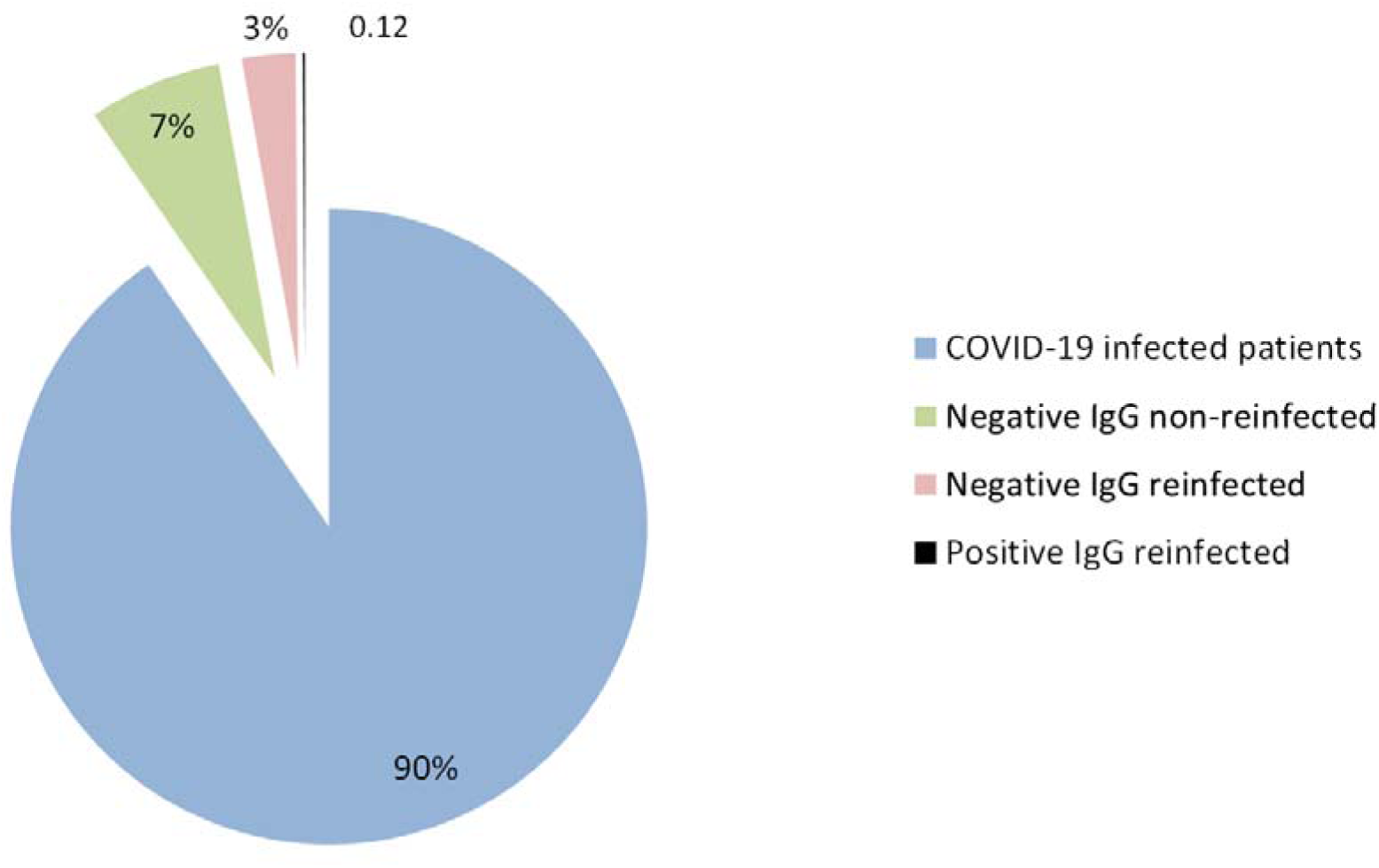
Patients with COVID-19. Among a total of 829 patients with COVID-19, 87 (10%) showed negative findings for IgG specific to SARS-CoV-2. Twenty-five (3%) patients were reinfected during the study period, while 62 (7%) patients remained healthy. A single patient with IgG positivity was reinfected.(0%)

Just after recovery, IgG antibodies against the SARS-CoV-2 were found in the serum of most of the reinfected patients. Only one patient was reinfected even though his IgG result remained positive after recovery from COVID-19 (Table 1). Most IgG-negative patients presented with just a couple of signs of COVID-19, including fever (96%) and myalgia (68%) and continued cough (< 15% cases), while reinfected patients suffered more signs including fever (96%), myalgia (88%), continuous cough (88%) and loss of taste and smell together (72%). In addition, after reinfection more than 95% of the reinfected COVID-19 patients had been immunised as evidenced by IgG antibody induction. Surprisingly, there was no detectable IgG concentration in a male patient who had most of the common signs and symptoms of COVID-19 during both his first infection and reinfection. Also, a male patient (no. 26 in Tables 1 and 2) showed serum IgG level of 5.87 s/ca against SARS-CoV-2 after recovery but was reinfected 138 days later. Interestingly, the reinfection induced his immune system to produce IgG level by amount (2.08 s/ca) less than the first infection. The occurrence of reinfection in the group ranged from 26 to 138 days after recovery from the initial infection (Table 2).

**Table 1:**
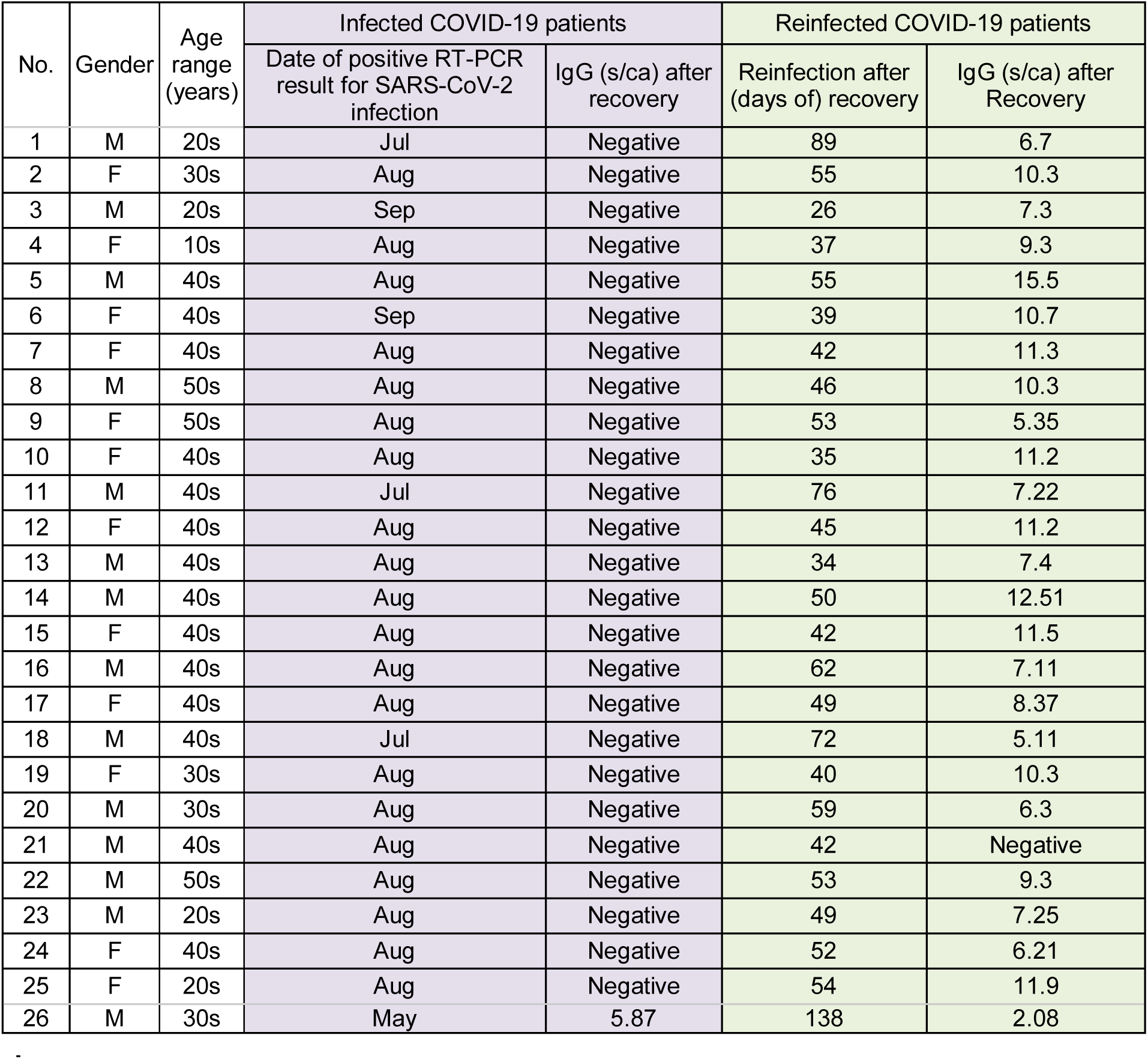
COVID-19 data of the 26 reinfected patients in this study

**Table 2:**
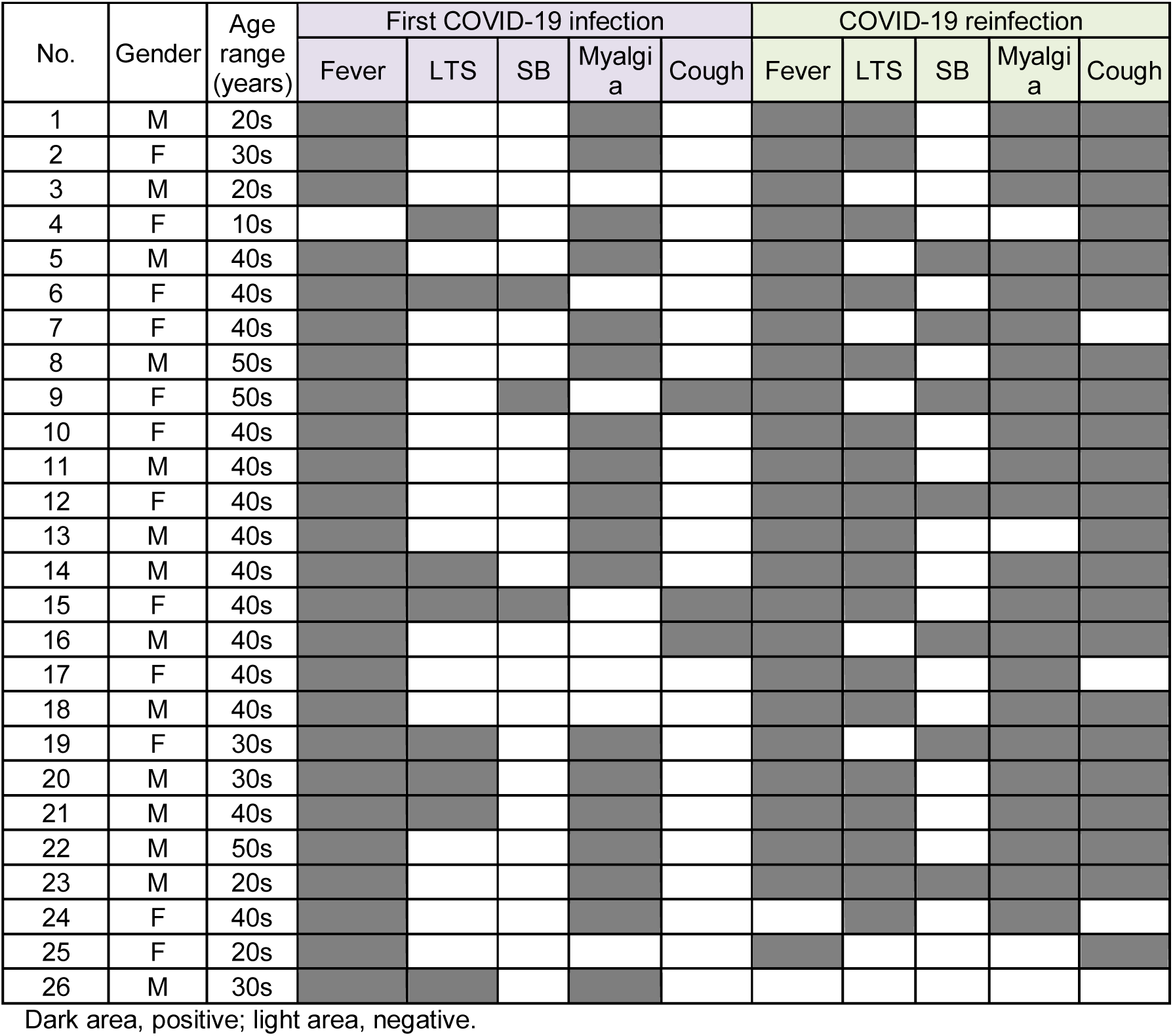
Sign and symptoms among all reinfected patients during both infection and reinfection

## Discussion

Approximately 90% of recovered COVID-19 patients produce a detectable level of IgG [25]. In our study, among 829 infected cases, 742 IgG-positive recovered patients were identified. The duration and viral magnitude can play a crucial role in inducing the immune system to produce an adequate IgG level [26-28], which can be an indicator of the severity of the disease [29]. Therefore, it may be postulated that those patients who recovered from SARS-CoV-2 infection with IgG negativity in this study were exposed to a lesser amount of viral antigen.

The degree of protective immunity conferred by prior infection and the possibility of reinfection by SARS-CoV-2 is not well understood [20]. This study reports that around 10% of recovered COVID-19 patients showed no detectable IgG concentration after recovery and prior to their reinfection. A pair of studies from Hong Kong and Ecuador have reported SARS-CoV-2 reinfection in two IgG-negative patients who previously recovered from COVID-19 [21, 22]. The IgG level appears to be associated with the severity of the illness during the first infection; studies have shown that patients with mild symptoms had no/lower antibody titers as compared with those patients with more severe symptoms [21, 29, 30]. In our study, the degree of disease severity in the reinfection period was worse in most patients than that during the first instance of COVID-19. A similar case study was reported in a 46-year-old male Ecuadorian patient [23]. This contradicts with the findings of a couple of case studies in which patients were asymptomatic during their reinfection period but were symptomatic during their first infection [21, 22]. The increase in disease severity in reinfected patients can be due to a high viral load or a change in virus virulence, which might have facilitated reinfection [20, 31]. The lack of a detectable level of IgG against SARS-CoV-2 during the infection period in mild or asymptomatic patients possibly makes them more susceptible to the reinfection. Therefore, in the current study, it is believed that the vast majority of patients who showed detectable levels of IgG after COVID-19 were thus protected from reinfection, even though the time period of the immunity conferred by IgG against SARS-CoV-2 has not been concluded yet [21]. Unexpectedly, a male patient was reinfected with COVID-19 without inducing IgG production a second time, which raises the question of possible reinfection for a third time. Also, another male patient had detectable amounts of IgG during his first infection and was reinfected after 138 days with no symptoms. That may be due to a decrease in his IgG level as time passed as studies have suggested neutralizing IgG levels start to decrease at six to 13 weeks after infection resolution [29, 32, 33]. Mysteriously, the reinfection in the aforementioned patient induced a low level of a detectable serum IgG concentration. This raises questions concerning the presence of adaptive immunity in COVID-19 patients.

## Conclusion

To conclude, a lack of IgG in patients who have recovered from COVID-19 may lead some to become infected. IgG production possibly indicates the severity of the signs and symptoms of COVID-19. Also, IgG levels against SARS-CoV-2 may decrease with time. Further studies are needed to consider the efficiency and sustainability of IgG, which are likely to play a vital role in the success of the COVID-19 vaccine industry.

## Data Availability

All data has been included with in the manuscript

## Author contributions

AMA performed lab work. KMA, MHF, HMT and HMR contributed to writing and preparing the manuscript.

## References

[1] Zhu N, Zhang D, Wang W, Li X, Yang B, Song J, et al. A Novel Coronavirus from Patients with Pneumonia in China, 2019. New England Journal of Medicine. 2020;382:727–33.

[2] Day M. Covid-19: four fifths of cases are asymptomatic, China figures indicate. BMJ. 2020;369:m1375.

[3] Bendavid E, Mulaney B, Sood N, Shah S, Ling E, Bromley-Dulfano R, et al. COVID-19 Antibody Seroprevalence in Santa Clara County, California. medRxiv. 2020:2020.04.14.20062463.

[4] Wu D, Wu T, Liu Q, Yang Z. The SARS-CoV-2 outbreak: What we know. International Journal of Infectious Diseases. 2020;94:44–8.

[5] Huang C, Wang Y, Li X, Ren L, Zhao J, Hu Y, et al. Clinical features of patients infected with 2019 novel coronavirus in Wuhan, China. The Lancet. 2020;395:497–506.

[6] Chen N, Zhou M, Dong X, Qu J, Gong F, Han Y, et al. Epidemiological and clinical characteristics of 99 cases of 2019 novel coronavirus pneumonia in Wuhan, China: a descriptive study. The Lancet. 2020;395:507–13.

[7] Chan JF-W, Yuan S, Kok K-H, To KK-W, Chu H, Yang J, et al. A familial cluster of pneumonia associated with the 2019 novel coronavirus indicating person-to-person transmission: a study of a family cluster. The Lancet. 2020;395:514–23.

[8] Yu P, Zhu J, Zhang Z, Han Y. A Familial Cluster of Infection Associated With the 2019 Novel Coronavirus Indicating Possible Person-to-Person Transmission During the Incubation Period. The Journal of Infectious Diseases. 2020;221:1757–61.

[9] Zou L, Ruan F, Huang M, Liang L, Huang H, Hong Z, et al. SARS-CoV-2 Viral Load in Upper Respiratory Specimens of Infected Patients. New England Journal of Medicine. 2020;382:1177–9.

[10] Rothe C, Schunk M, Sothmann P, Bretzel G, Froeschl G, Wallrauch C, et al. Transmission of 2019- nCoV Infection from an Asymptomatic Contact in Germany. New England Journal of Medicine. 2020;382:970–1.

[11] Ju B, Zhang Q, Ge J, Wang R, Sun J, Ge X, et al. Human neutralizing antibodies elicited by SARS- CoV-2 infection. Nature. 2020;584:115–9.

[12] Ravioli S, Ochsner H, Lindner G. Reactivation of COVID-19 pneumonia: A report of two cases. Journal of Infection. 2020;81:e72–e3.

[13] Felsenstein S, Herbert JA, McNamara PS, Hedrich CM. COVID-19: Immunology and treatment options. Clinical Immunology. 2020;215:108448.

[14] Clohisey S, Baillie JK. Host susceptibility to severe influenza A virus infection. Critical Care. 2019;23:303.

[15] Nogales A, L DeDiego M. Host single nucleotide polymorphisms modulating influenza A virus disease in humans. Pathogens. 2019;8:168.

[16] Iyer AS, Jones FK, Nodoushani A, Kelly M, Becker M, Slater D, et al. Persistence and decay of human antibody responses to the receptor binding domain of SARS-CoV-2 spike protein in COVID-19 patients. Science Immunology. 2020;5:eabe0367.

[17] Isho B, Abe KT, Zuo M, Jamal AJ, Rathod B, Wang JH, et al. Persistence of serum and saliva antibody responses to SARS-CoV-2 spike antigens in COVID-19 patients. Science Immunology. 2020;5:eabe5511.

[18] Chen D, Xu W, Lei Z, Huang Z, Liu J, Gao Z, et al. Recurrence of positive SARS-CoV-2 RNA in COVID-19: A case report. International Journal of Infectious Diseases. 2020;93:297–9.

[19] Xiao AT, Tong YX, Gao C, Zhu L, Zhang YJ, Zhang S. Dynamic profile of RT-PCR findings from 301 COVID-19 patients in Wuhan, China: A descriptive study. Journal of Clinical Virology. 2020;127:104346.

[20] Tillett RL, Sevinsky JR, Hartley PD, Kerwin H, Crawford N, Gorzalski A, et al. Genomic evidence for reinfection with SARS-CoV-2: a case study. The Lancet Infectious Diseases.

[21] To KK-W, Hung IF-N, Ip JD, Chu AW-H, Chan W-M, Tam AR, et al. Coronavirus Disease 2019 (COVID-19) Re-infection by a Phylogenetically Distinct Severe Acute Respiratory Syndrome Coronavirus 2 Strain Confirmed by Whole Genome Sequencing. Clinical Infectious Diseases. 2020.

[22] Van Elslande J, Vermeersch P, Vandervoort K, Wawina-Bokalanga T, Vanmechelen B, Wollants E, et al. Symptomatic SARS-CoV-2 reinfection by a phylogenetically distinct strain. Clinical Infectious Diseases. 2020.

[23] Prado-Vivar B, Becerra-Wong M, Guadalupe JJ, Marquez S, Gutierrez B, Rojas-Silva P, et al. COVID-19 Re-Infection by a Phylogenetically Distinct SARS-CoV-2 Variant, First Confirmed Event in South America. First Confirmed Event in South America(September 3, 2020). 2020.

[24] Ali KM, Ali AM, Tawfeeq HM, Figueredo G, Rostam HM. Hypoalbuminemia in Severe COVID-19 Post Recovery Patients. 2020.

[25] ECDC. Reinfection with SARS-CoV-2: considerations for public health response. European Centre for Disease Prevention and Control; 2020.

[26] Liu Y, Yan L-M, Wan L, Xiang T-X, Le A, Liu J-M, et al. Viral dynamics in mild and severe cases of COVID-19. The Lancet Infectious Diseases. 2020;20:656–7.

[27] Zhou F, Yu T, Du R, Fan G, Liu Y, Liu Z, et al. Clinical course and risk factors for mortality of adult inpatients with COVID-19 in Wuhan, China: a retrospective cohort study. The Lancet. 2020;395:1054–62.

[28] Fourati S, Hue S, Pawlotsky J-M, Mekontso-Dessap A, de Prost N. SARS-CoV-2 viral loads and serum IgA/IgG immune responses in critically ill COVID-19 patients. Intensive Care Medicine. 2020;46:1781–3.

[29] Long Q-X, Tang X-J, Shi Q-L, Li Q, Deng H-J, Yuan J, et al. Clinical and immunological assessment of asymptomatic SARS-CoV-2 infections. Nature Medicine. 2020;26:1200–4.

[30] Robbiani DF, Gaebler C, Muecksch F, Lorenzi JCC, Wang Z, Cho A, et al. Convergent antibody responses to SARS-CoV-2 in convalescent individuals. Nature. 2020;584:437–42.

[31] Guallar MP, Meiriño R, Donat-Vargas C, Corral O, Jouvé N, Soriano V. Inoculum at the time of SARS-CoV-2 exposure and risk of disease severity. International Journal of Infectious Diseases. 2020;97:290–2.

[32] Wang X, Guo X, Xin Q, Pan Y, Li J, Chu Y, et al. Neutralizing Antibodies Responses to SARS-CoV-2 in COVID-19 Inpatients and Convalescent Patients. medRxiv. 2020:2020.04.15.20065623.

[33] Kissler SM, Tedijanto C, Goldstein E, Grad YH, Lipsitch M. Projecting the transmission dynamics of SARS-CoV-2 through the postpandemic period. Science. 2020;368:860–8.

